# Market Surveillance of the Microbial Quality of Smoked Fish in the Sekyere Enclave of the Ashanti Region, Ghana

**DOI:** 10.1101/2025.03.21.25324371

**Authors:** Denis Dekugmen Yar, William K. J. Kwenin, Doreen Dedo Adi, Ebenezer Assoah, Simon Nyarko, Gadafi Iddrisu Balali, Augustine Dadzie

## Abstract

This study assessed the microbial quality of selected smoked fish: *Thunnus spp*.[tuna], *Oreochromis niloticus* [tilapia], *Clarias gariepinus* [catfish], *Clupea harengus* [herring], and *Salmo salar* [salmon] commonly sold and consumed in the main markets of the Sekyere Enclave of the Ashanti Region, Ghana. Sixty smoked fish were sampled from these sites, Mampong, Kofiase, Agona, and Nsuta markets. The samples were then processed and analyzed using rigorous standard microbiological techniques and assays to determine viable bacterial counts and isolates. The means Aerobic Plate Count for the smoked fish samples were 1.47±0.012×10^5^, 3.558±0.11×10^4^, 1.854±0.02×10^4^ and 1.038±0.01×10^4^ (cfu/g) for the Kofiase, Agona, Mampong, and Nsuta market, respectively, with an overall mean of 2.115 ±0.23×10^5^cfu/g. Microbial loads (cfu/g) for Salmon, Tuna, Catfish, Herring, and Tilapia were 4.69±0.11×10^5^, 3.63±0.021×10^5^, 2.29±0.12×10^5,^ 1.93±0.11×10^5^, and 9.0±0.04×10^4^, respectively, with an overall mean count of 1.34±0.13×10^6^cfu/g. Seven different bacteria genera were isolated: *Klebsiella* spp (18.24%), *E*. *coli* (16.98%), *Vibrio* spp. (15.09%), *Pseudomonas* spp. (14.47%), *Staphylococcus* spp (13.21%), *Shigella* spp (11.32%), and *Salmonella* spp (10.69%). This study’s results demonstrate that smoked fish were heavily contaminated with potential bacterial pathogens and could cause severe foodborne diseases. Therefore, immediate and strict adherence to food hygiene and safety standards should be enforced to avoid potential foodborne outbreaks in the Enclave.

**Author Summary:** This study evaluates the microbial quality of smoked fish sold in markets across the Sekyere Enclave, Ashanti Region, Ghana. Sixty samples of five fish species; tuna, tilapia, catfish, herring, and salmon were analyzed using standard microbiological methods. The findings revealed high microbial loads, with an overall mean of 1.34×10⁶ cfu/g. Seven bacterial genera were identified, including *Klebsiella* spp., *E. coli*, and *Salmonella* spp., indicating significant contamination. These results highlight smoked fish as a potential source of foodborne diseases due to improper handling and storage practices. Given the widespread consumption of smoked fish in the region, this study highlights the need for improved food safety practices and regulatory measures to minimize health risks. By identifying critical risk factors, the study contributes to safeguarding public health and enhancing food hygiene standards in Ghana.

## 1. Introduction

Fish is a significant source of protein, vitamins, and minerals that supplements our diets and contains 10% of calories [1] and more than 60% of protein intake in the adult population [2, 3]. Fishes are a source of food, provide recreational opportunities, and play a significant role in trade [2–4]. In addition, fishes offer numerous benefits to humans, spanning across various aspects of life such as culture, which includes spiritual elements like sacred and religious practices, and inspirational aspects like aesthetic art and folklore [2–4].

Fish, however, is highly perishable by nature, and many strategies have been developed over the years to limit its spoilage [5]. In Ghana, various preservation techniques are utilized to extend the shelf life of fish and prevent spoilage. These methods include smoking, salting, frying, sun-drying, freezing, and fermentation [6]. Smoking fish with wood preserves and extends its shelf life through its antibacterial and oxidative properties and enhances its flavour profile and aroma, making it more appealing for consumption [7, 8].

The Sekyere Enclave is a market hub for smoked fish in the region because of its proximity to the tributaries of the Volta Lake, where fishing is the primary occupation of the people [9]. Smoked fish products from the lake sites are subsequently brought and distributed to markets on wholesale or retail bases, exposing them to microbial contaminations. These smoked fish products are stored at room temperature in the marketplaces for weeks and displayed for sale in the open markets, making them susceptible to contamination [10]. Consumers of smoked fish are, therefore, prone to health hazards due to possible microbial contamination [11].

Data on the risk factors for foodborne diseases indicate that most outbreaks result from inappropriate food handling practices, marketing, storage, and, for smoked fish products, partial removal of water during production [12, 13]. However, there are few studies on market surveillance of the microbial quality of smoked fish in the Sekyere Enclave and its environs, which could be contributing to the increasing cases of foodborne-related illnesses [14, 15]. Meanwhile, Smoked fish is highly consumed in this study area because it is available and affordable for most residents. However, limited literature examined the microbial profile of smoked fish and its link with foodborne illnesses in this area. Thus, this calls for a concerted effort to examine the microbial quality of smoked fish as a possible risk factor for foodborne illness in this area. This study, therefore, profiled the microbial quality of some commonly marketed smoked fish using indicator bacteria.

## 2. Materials and Methods

### Design of the study

This study employed a market-based cross-sectional survey to determine the microbial quality of commonly marketed and consumed smoked fish in the Sekyere Enclave of the Ashanti Region.

### Study Areas

This study was conducted in the Asante-Mampong, Kofiase, Agona, and Nsuta markets of the Sekyere Enclave of the Asante Region of Ghana. These districts and town markets are near Yeji, a fishing hub where fish traders from the Sekyere Enclave buy smoked or fresh fish for sale within the Enclave [16]. In these communities, there are designated areas for smoked fish vending, especially on market days, although some vendors also sell informally along roadsides.

### Mampong

Mampong is the municipality capital located in the northern part of the region, about 57km from the regional capital, Kumasi, and has a total landmass of approximately 449 km^2^ [17]. The municipality is bounded on the south by Sekyere South District, the east by Sekyere Central, and the north by Ejura-Sekyere Dumase Municipal. It has 79 settlements, about 61% rural and 31% forming the urban enclave. The population of the municipality is 88,051, with a growth rate of 1.8% annually [18]. It has an average annual rainfall of 1270mm and two rainy seasons [19].

### Kofiase

Kofiase is one of the sub-districts of the Asante-Mampong Municipal, 5km from the municipal capital and 52 km from Kumasi. It is rural and has a significant market for smoked fish business [17].

### Agona

Agona is the capital of the Sekyere South District in the region. It is located in the eastern part of the region and has a total land area of 770 km^2^. The population is 116,477, and the growth rate is 2.2% annually [20]. The geographic coordinates for Agona are Latitude 6°47’N and Longitude 1°36’W [20].

### Nsuta

Nsuta is the capital of the Sekyere Central District and is located in the northern part of the region. The geographic coordinates for Nsuta are Latitude 7°00′40″N and Longitude 1°22′53″W. It has a total land size of 2,345 km^2^, with a population of 73,228 [18].

### Study Population and Sample Size

The study was conducted on smoked fish obtained from fish traders, predominantly females aged 18-70 with little formal education. Sixty smoked fish were estimated for the study, 15 samples per market. The study used five different species of fish: *Clupea harengus* (herring) and *Thunnus* spp. (tuna), *Clarias gariepinus* (catfish), *Salmo salar* (salmon), and *Oreochromis niloticus* (tilapia). Three samples of each fish species from each market are summarized in Table 1.

**Table 1:** Fish Samples from the Selected Study Areas.

| Site/Fish | *Thunnus*<br>*spp.* | *Clupea*<br>*harengus* | *Salmo*<br>*salar* | *Oreochromis*<br>*niloticus* | *Clarias*<br>*gariepinus* | Total |
| --- | --- | --- | --- | --- | --- | --- |
| Mampong | 3 | 3 | 3 | 3 | 3 | 15 |
| Agona | 3 | 3 | 3 | 3 | 3 | 15 |
| Nsuta | 3 | 3 | 3 | 3 | 3 | 15 |
| Kofiase | 3 | 3 | 3 | 3 | 3 | 15 |
| Total | 12 | 12 | 12 | 12 | 12 | 60 |

(*Thunnus* spp.[Tuna], *Oreochromis niloticus* [Tilapia], *Clarias gariepinus* [Catfish], *Clupea harengus* [Herring], and *Salmo salar* [Salmon]).

### Sampling techniques

The four study sites were selected purposively based on their closeness to Yeji, a well-known fishing community, the availability of fishing markets, and the level of fish trading business. A systematic sampling technique was employed to collect smoked fish using aseptic techniques into sterile zip bags. The smoked fish samples were obtained through buying from the sellers, placed in ice packs, and quickly transported to the Microbiology Laboratory (AAMUSTED-M) for analysis.

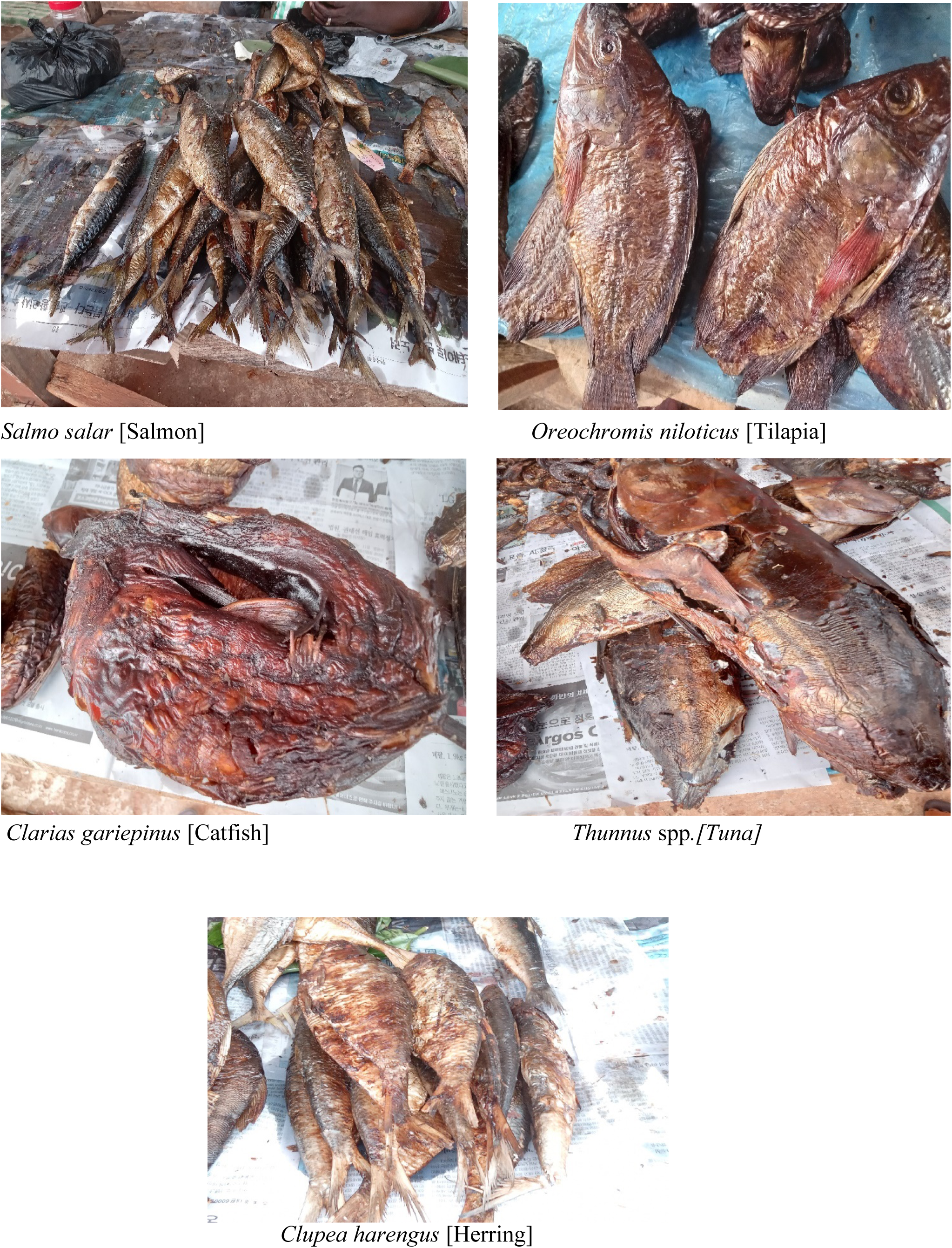

### Laboratory Methods and Analysis

#### Media Preparation

MacConkey Agar (MCA), Mannitol Salt Agar (MSA), Cetrimide Agar (CA), Salmonella-Shigella Agar (SSA), Thiosulfate Citrate Salt Bile Sucrose Agar (TCSBS), Nutrient Agar (NA), Aerobic Plate Count Agar (APCA), and others were prepared based on the manufacturers’ standard protocols and procedures, as previously reported [15, 21].

#### Preparation of Fish Samples

Each smoked fish sample weighed 25 g, was added to 225 ml of Phosphate Buffer Solution (PBS), and stomached in a sterile Drawell Lab Stomacher blender (Model: DW-04, China). A tenfold serial dilution of each sample was made and used for the bacteria culture.

#### Culture and Isolation

Triplicates of a 1ml of the 10^-4^ serial dilution each were pipetted and plated on the following media: SSA, MSA, TCSBS, CA, and APCA for isolation and differentiation of the various bacterial groups: *Salmonella* and *Shigella* spp., *Staphylococcus* spp., *Vibrio* spp., *Pseudomonas* spp., and total aerobic counts, respectively, as previously described (Yar et al., 2021). The plates were then incubated at 35°C for 24 hours. Viable microbial load counts were achieved by counting discrete colonies using the colony counter (Stuart ® Model: SC6, UK). The microbes were identified using standard microbiological methods and biochemical processes, as previously reported [15, 21, 22].

#### Identification of Bacteria

Colourless colonies with or without black centres and pink with pink zones on the SSA plates indicated the presence of *Salmonella* and *Shigella* spp, respectively. Cream and yellow to golden colonies on MSA were identified as *Staphylococcus* spp. Bacteria isolated on CA medium were suggestive of *Pseudomonas* spp. Pink to red mucoid colonies on the MacConkey medium were suggestive of *Klebsiella* spp, while pink to red non-mucoid colonies were suggestive of *E. coli.* These isolates were differentiated as previously described [15, 21, 22]. Yellow colonies on the surface of the TCSBS medium were suggestive of *V. cholerae,* and black colonies of *V. parahaemolyticus* and differentiated as previously described [21].

### Statistical Analysis

The raw data from the microbial analysis were compiled using a Microsoft Excel 2021 spreadsheet. The counts were then transformed into log_10_. Using STATISTIX 9.0 (Analytical Software, 2008), descriptive statistics analysis (mean, percentages, frequency) was performed, and the results were presented in tables and graphs.

### Ethical Consideration

Before data collection, formal written permission was obtained from the Environmental Health Directorates. Fish samples were bought from individual vendors; hence, no written permission was necessary.

## 3. Results

Table 2 shows the mean Aerobic Plate Count (APC) of smoked fish at the four study markets measured in cfu/g. The total microbial mean counts in smoked fish sampled from the different markets were 1.47±0.012×10^5^, 3.558±0.11×10^4^, 1.854±0.02×10^4^ and 1.038±0.01×10^4^ (cfu/g) from the Kofiase, Agona, Mampong, and Nsuta market, respectively, with an overall mean APC of 2.115×10^5^ cfu/g.

**Table 2.** Mean Microbial Counts of smoked fish in Mampong, Agona, Nsuta, and Kofiase.

| Area | Sample Size | Aerobic Plate Count (cfu/g) | Mean log (cfu/g) |
| --- | --- | --- | --- |
| Mampong | 15 | $1.854 \pm 0.02 \times 10^4$ | 4.268 |
| Agona | 15 | $3.558 \pm 0.11 \times 10^4$ | 4.551 |
| Nsuta | 15 | $1.038 \pm 0.01 \times 10^4$ | 4.016 |
| Kofiase | 15 | $1.47 \pm 0.012 \times 10^5$ | 5.167 |
| **Total** | **60** | **$2.115 \pm 0.23 \times 10^5$** | **5.325** |

**Table 3** shows the average microbial loads of the five smoked fish sampled: *Salmo salar* [Salmon], *Thunnus spp.* [Tuna], *Clarias gariepinus*[catfish], *Clupea harengus*[Herring], and *Oreochromis niloticus*[Tilapia] which yielded microbial counts of 4.69 ± 0.11×10^5^, 3.63 ± 0.021×10^5^, 2.29±0.12×10^5,^ 1.93 ± 0.11×10^5^cfu/g, and 9.0 ± 0.04×10^4^cfu/g, respectively with overall total mean count of 1.34 ± 0.13×10^6^ (cfu/g).

**Table 3.** Mean Microbial Counts in Smoked Fish sampled from the different Markets.

| Fish | Sample size | Total mean count (cfu/g) | Mean log (cfu/g) |
| --- | --- | --- | --- |
| *Thunnus* spp. [tuna] | 12 | 3.63 ± 0.021×10<sup>5</sup> | 5.559 |
| *Salmo salar* [salmon] | 12 | 4.69 ± 0.11×10<sup>5</sup> | 5.671 |
| *Oreochromis niloticus* [tilapia] | 12 | 9.0 ± 0.04×10<sup>4</sup> | 4.954 |
| *Clupea harengus* [herring] | 12 | 1.93 ± 0.11×10<sup>5</sup> | 5.285 |
| *Clarias gariepinus*[catfish] | 12 | 2.29 ± 0.12 ×10<sup>5</sup> | 5.359 |
| **Total** | **60** | **1.34 ± 0.13×10<sup>6</sup>** | **6.128** |

**Table 4** summarizes the prevalence of bacteria isolated from smoked fish across the different study markets, with rates of 49.05%, 22.64%, 14.47%, and 13.84% observed in Kofiase, Agona, Mampong and Nsuta, respectively. Figure 2 and **Table 5** show the proportions of bacteria species identified among the smoked fish: *Klebsiella* spp. (18.24%), *E. coli* (16.98%), *Vibrio* spp. (15.09%), *Pseudomonas* spp. (14.47%), *Staphylococcus* spp. (13.21%), *Shigella* spp. (11.32%), *Salmonella* spp. (10.69%). Additionally, **Table 5** shows the bacteria species distribution among the different smoked fish species: *Thunnus* spp. (23.90%), *Salmo salar* (27.04%), *Oreochromis niloticus* (6.92%), *Clupea harengus* (20.76%), and *Clarias gariepinus* (21.38%).

**Figure 1.**
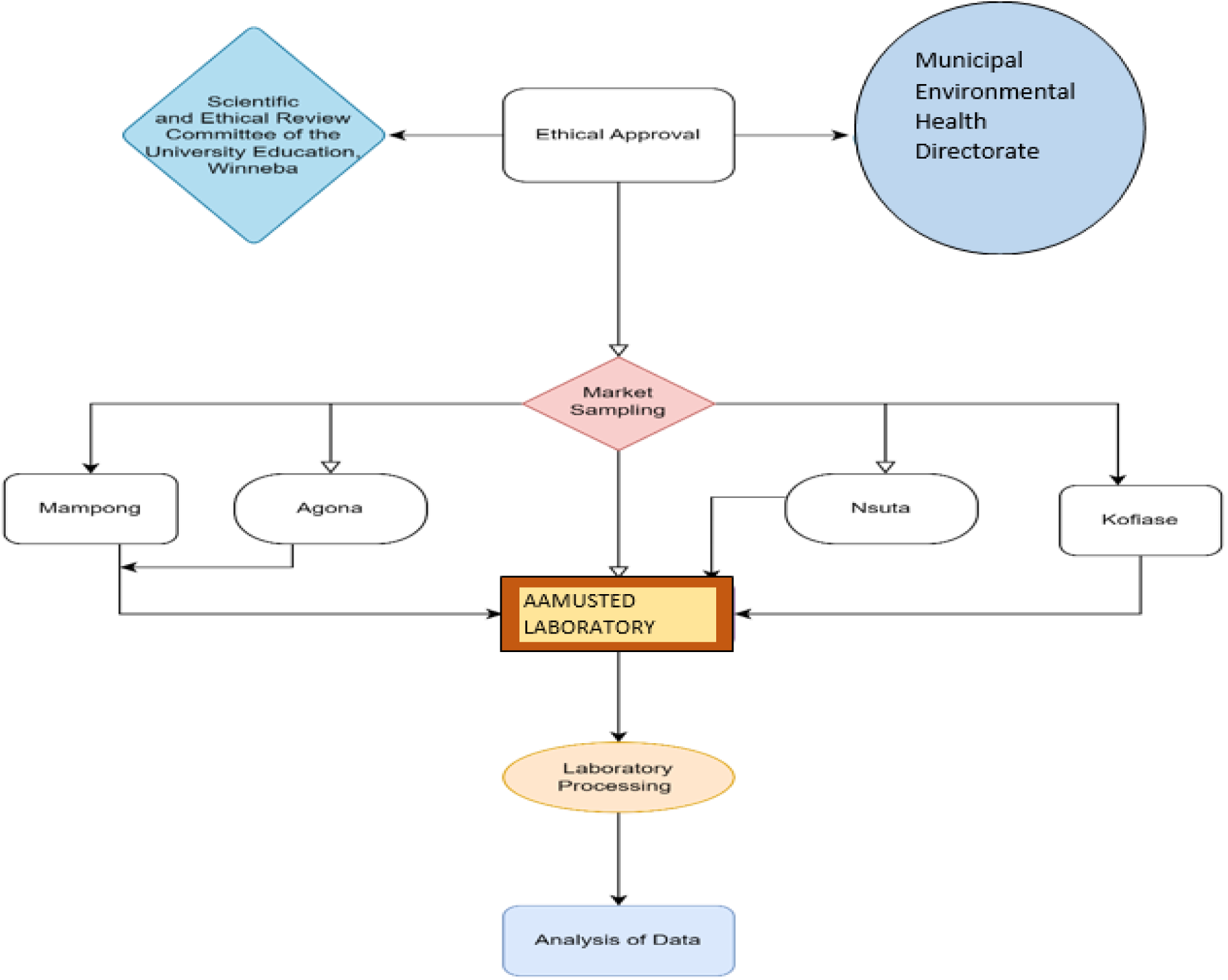
Schematic diagram of the workflow.

**Figure 2:**
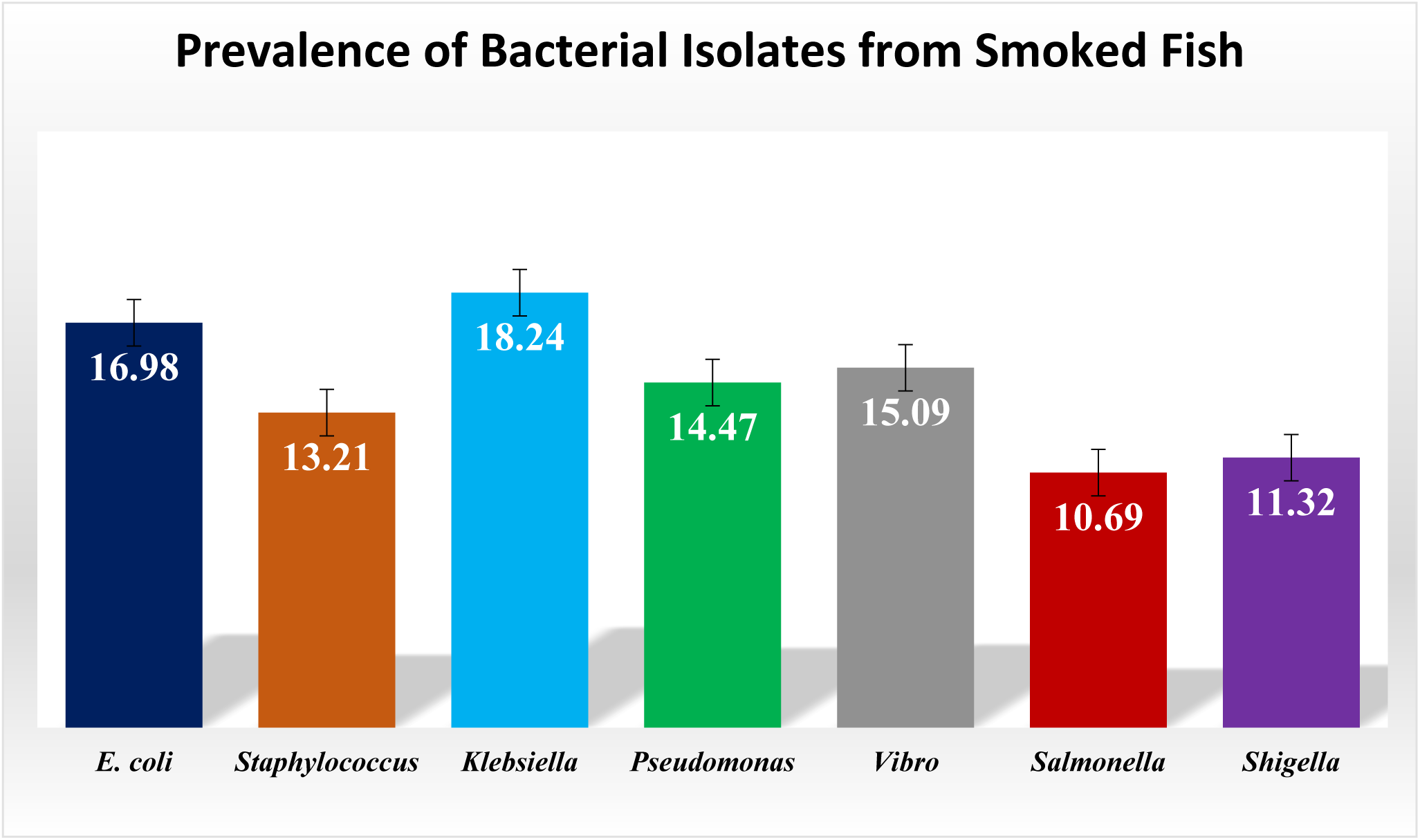
Proportions of Bacteria isolated from Smoked Fish in the Sekyere Enclave.

**Table 4:** Bacteria Isolated in Smoked Fish from Markets in the Study Area.

| Study Site | *E. coli* (%) | *Staph.* (%) | *Kleb.* (%) | *Pseud* (%) | *Vibrio* (%) | *Salm.* (%) | *Shigella* (%) | Total (%) |
| --- | --- | --- | --- | --- | --- | --- | --- | --- |
| Mampong | 3<br>(11.11) | 4<br>(19.05) | 3<br>(10.34) | 1<br>(0.04) | 3<br>(0.13) | 4<br>(23.53) | 5<br>(27.78) | 23<br>(14.47) |
| Agona | 4<br>(14.81) | 5<br>(23.81) | 6<br>(20.69) | 4<br>(17.39) | 4<br>(16.67) | 7<br>(41.18) | 6<br>(33.33) | 36<br>(22.64) |
| Nsuta | 5<br>(18.52) | 0<br>(0) | 5<br>(17.24) | 7<br>(30.43) | 5<br>(20.83) | 0<br>(0) | 0<br>(0) | 22<br>(13.84) |
| Kofiase | 15<br>(55.55) | 12<br>(57.14) | 15<br>(51.72) | 11<br>(47.83) | 12<br>(50.0) | 6<br>(35.29) | 7<br>(41.18) | 78<br>(49.05) |
| **Total** | **27**<br>**(16.98)** | **21**<br>**(13.21)** | **29**<br>**(18.24)** | **23**<br>**(14.47)** | **24**<br>**(15.09)** | **17**<br>**(10.69)** | **18**<br>**(11.32)** | **159**<br>**(100)** |

*E. coli*-*Escherichia coli*, *Staph*-*Staphylococcus* spp., *Kleb.*- *Klebsiella* spp, *Pseud.*- *Pseudomonas* spp., *Vibrio*-*Vibrio* spp., *Salm.*-*Salmonella* spp.

**Table 5.** Microbial Isolates from different Smoked Fishes sampled from the Study Sites.

| Type of Fish | *E. coli*<br>(%) | *Staph.*<br>(%) | *Kleb.*<br>(%) | *Pseudo*<br>(%) | *Vibro*<br>(%) | *Salm.*<br>(%) | *Shigella.*<br>(%) | Total<br>(%) |
| --- | --- | --- | --- | --- | --- | --- | --- | --- |
| *Thunnus spp* | 5<br>(18.52) | 6<br>(28.57) | 5<br>(17.24) | 8<br>(34.78) | 4<br>(16.67) | 5<br>(29.41) | 5<br>(27.77) | 38<br>(23.90) |
| *Salmo salar* | 7<br>(25.93) | 6<br>(28.57) | 7<br>(24.14) | 5<br>(21.74) | 7<br>(29.16) | 5<br>(29.41) | 6<br>(33.33) | 43<br>(27.04) |
| *Oreochromis niloticus* | 4<br>(14.81) | 0<br>(0.00) | 4<br>(13.79) | 1<br>(4.35) | 0<br>(0.00) | 1<br>(5.88) | 1<br>(5.56) | 11<br>(6.92) |
| *Clupea harengus* | 6<br>(22.22) | 5<br>(23.81) | 6<br>(20.69) | 4<br>(17.39) | 6<br>(25.00) | 3<br>(17.65) | 3<br>(16.67) | 33<br>(20.76) |
| *Clarias gariepinus* | 5<br>(18.52) | 4<br>(19.05) | 7<br>(24.14) | 5<br>(21.74) | 7<br>(29.17) | 3<br>(17.65) | 3<br>(16.67) | 34<br>(21.38) |
| **Total** | **27**<br>**(16.98)** | **21**<br>**(13.21)** | **29**<br>**(18.24)** | **23**<br>**(14.47)** | **24**<br>**(15.09)** | **17**<br>**(10.69)** | **18**<br>**(11.32)** | **159**<br>**(100)** |

*E. coil*-*Escherichia coli*, *Staph* - *Staphylococcus*, *Kleb.* – *Klebsiella*, *Pseud.*- *Pseudomonas*, *Vibrio*-*Vibrio spp*, *Salm.* -*Salmonella*

## 4 Discussions

### Microbial contaminations of smoked fish per markets

This study used standard microbiological techniques to examine the microbial quality of commonly marketed and consumed smoked fish in the Sekyere Enclave of the Ashanti Region. We reported an average microbial load of >10^4^/g from all markets surveyed, higher than the permissible limits of <10^4^/g in smoked fish or food products set by the Ghana Standard Authority [15], the Food and Agricultural Organization [23], and Public Health Ontario [24]. The use of total plate count is a tool for monitoring meat quality and the level of contamination. This study validates that smoked fish sold in these markets were heavily colonized with bacteria and could pose a health risk to consumers.

The high bacterial loads reported in this study could be due to poor smoking, improper storage conditions, or post-smoked processing temperature abuse [25]. The high loads may also result from exposure of smoked fish to environmental contaminants [24]. Furthermore, poor adherence to food safety and hygienic practices and little food safety knowledge among smoked fish vendors, coupled with storage facilities, transportation, and market governance, could affect the quality and safety of smoked fish [26]. Additionally, in these markets, vendors would often display smoked fish on table tops or the floor, exposing them to dust and insects [10], compromising the microbial quality.

In this study, smoked fish from the Kofiase market were significantly colonized with bacteria compared to other markets. Markets with stalls and concrete floors had limited particulate dust and adequate sanitation with few environmental hazards exposure to the smoked fish. The highest bacteria counts found in the Kofiase market could be attributed to the microbial quality of the market environment [27]. It is, however, worth noting that these markets operate outdoors, with vendors displaying goods in the open, exposing them to environmental contaminants [10].

This study observed a possible link between market hygiene and the microbial quality of the smoked fish. The traders in these markets hardly adhere to food safety standards, such as wearing head mesh, aprons and gloves while handling fish. Conversely, some vendors exhibited superior hygiene practices by storing smoked fish in baskets covered with brown sheets and displaying them on table tops covered with polyethene [27]. This practice could have contributed to the relatively low microbial loads and isolate observed in the Nsuta and Mampong markets. The behaviour of consumers and sellers at these markets could equally have contributed to the microbial abundance due to their frequent handling and touching of the fish with bare hands in an attempt to buy and sell [27].

### Microbial contaminations per the smoked fish species

In this study, the mean microbial counts of at least >10^4^/g in all five fish species were higher than the accepted standard indicated earlier. This study shows that smoked *Salmo salar* [salmon], *Thunnus spp.* [tuna], *Clarias gariepinus* [catfish], *Clupea harengus* [herring], and *Oreochromis niloticus* [tilapia] were highly colonized by microorganisms, and this aligns with studies in Ghana [28, 29]. Smoked salmon was more colonized with bacteria than the other fish species [28]. The disparity of bacteria load on the different fish species may be attributed to intrinsic factors such as pH, moisture, and the nutrient contents of the fish species [30]. Smoked salmon generally has high moisture and protein contents and can support microbial growth compared to the other species [31]. Conversely, Tilapia has low moisture and protein contents to support bacterial growth and thus recorded the lowest load in this study.

### Microbial Isolates per Markets

In this study, *Klebsiella* spp., *E. coli*, *Vibrio* spp., *Pseudomonas* spp., *Staphylococcus* spp., *Shigella* spp., and *Salmonella* spp., were isolated from smoked fish sampled in these markets, except in Nstuta. Previous studies in Ghana recovered these bacteria among smoked fish [29, 32]. However, another comparable study isolated similar bacteria among smoked fish in addition to *Enterobacter* spp., *Campylobacter* spp., and *Listeria monocytogenes* [32]. The different species of bacteria recovered among the smoked fish in these studies could be linked to several factors, including sources of the fish harvested, fish processes, smoking methods, storage, packaging, transportation, and laboratory approaches [33].

This study recovered *E. coli* and *Staphylococcus* spp among the smoked fish in most markets and measured vendor hygiene standards. It must, however, be noted that fish in their natural environments harbour various types of bacteria species due to constant exposure to contaminated water. These bacteria, therefore, may result from direct or indirect contamination of the fish [12, 13, 26].

*Klebsiella* spp and *Pseudomonas* spp indicate the level of spoilage [34]. In contrast, *Vibrio* spp, *Shigella* spp, and *Salmonella* spp isolated suggest food safety standards among sellers [15]. The presence of these pathogenic bacteria in smoked fish poses risks of foodborne diseases. Therefore, smoked fish in the Sekyere enclave could be unwholesome for consumers if not cooked before eating.

In this study, *Klebsiella* spp was the predominant bacteria isolated at 18%, higher than the 7% in a similar study in Nigeria [34], but lower than the reported prevalence rate by Adesoji et al. (2019). The differences may result from poor sanitary conditions in the different markets and inadequate adherence to hygiene practices among the smoked fish vendors [35].

*Salmonella* spp. was the lowest prevalent isolated (11%), somewhat similar to the 14% [29] and lower than the 67% [36] reported in northern Ghana. This current study, however, disagrees with a study in Ghana that did not find *Salmonella* spp. in smoked fish [28]. Kofiase recorded the highest prevalence of isolates, which indicates the markets’ states, the level of community development, and vendors’ adherence to food hygiene and safety standards.

## Conclusion

This study shows that the microbial loads among smoked fish in the Sekyere Enclave exceeded the minimum permissible levels recommended by local and international food regulators. Smoked fish were heavily contaminated with pathogenic bacteria, especially in rural markets, compromising their quality and posing a health threat to consumers. Therefore, it is recommended that vendors should be trained on food safety and hygiene standards to improve fish’s microbial quality and avoid possible food-borne outbreaks in the Enclave.

## Data Availability

All relevant data are within the manuscript and its Supporting Information files.

## Acknowledgements

We sincerely acknowledge the effort of Godfred Mensah, Jennifer Mensah, Gideon Nimfour, Elvis Osei Tutu, Elizabeth Eduam, Dennis Atakora Amaniampong, former students of the Akenten Appiah-Menka University of Skills Training and Entrepreneurial Development (AAMUSTED), Asante Mampong Campus. Their dedication to data collection during their voluntary attachment at the health facilities was instrumental to this study.

## Availability of supporting data

All data used for this manuscript are available upon a reasonable request.

## Authors Contributions

DDY played a pivotal role in conceptualizing and developing the manuscript, provided critical edits, contributed to data analysis and ensured the manuscript’s intellectual rigor and clarity. WKJK participated in data analysis and presentation, reviewed the manuscript and approved it for publication. DDA reviewed the manuscript and approved it for publication. EA participated in data collection, data analysis and presentation, and drafting of the manuscript. SN participated in drafting of the manuscript and reviewed the manuscript for publication. GIB reviewed and structured the manuscript for publication. AD responsible for data collection and reviewed the manuscript. All authors have thoroughly read and approved the final manuscript for publication.

## Consent for publication

We (authors) have all read the final manuscript and consented to it for publication.

